# Bacterial profile and antibiotic sensitivity among patients admitted to the Intensive Care Unit at Mbeya Zonal Referral Hospital: a cross-sectional study

**DOI:** 10.64898/2026.10.14.26365595

**Authors:** M.A. Zumba, S. Mrutu, A. Jongo, E. Lugazia, H. Msanga, A. Omari, F. Ringo, M. Elias

## Abstract

Sepsis is a global health problem. Patients in the Intensive Care Unit (ICU) are at risk of having sepsis because of their vulnerability and use of invasive medical devices. Emerging antimicrobial-resistance complicates treatment. Understanding bacterial profiles and antibiotic sensitivity patterns in the ICU plays an important role in managing sepsis, optimizing antimicrobial use, and improving patient outcomes. The aim of this study was to show bacterial profiles and antibiotic sensitivity patterns among patients who were admitted to the ICU at Mbeya Zonal Referral Hospital for 12 months. This was a prospective cross-sectional study among 187 sepsis-suspected patients. Data analysis was performed using SPSS Version 27. Categorical data were summarized by frequency and percentage. Fisher’s exact test, chi-square test, and modified Poisson regression were used to show the association between patients’ clinical variables and culture positivity at a 95% CI. Statistical significance was set at a p-value < 0.05. Out of 187 samples, 41.7% were culture positive. Gram-negative bacteria were commonly isolated (65.4%, with Klebsiella pneumoniae and Escherichia coli being commonly isolated. Commonly isolated Gram-positive bacteria were Coagulase-negative Staphylococcus species (19%). 100% of gram-positive bacteria and 96% of gram-negative bacteria were Multi-Drug Resistant (MDR), respectively. In multivariable analysis, patients who were intubated and mechanically ventilated (aPR = 2.208, 95% CI: 1.133-4.305, p = 0.020) and post-laparotomy patients (aPR = 3.241,95% CI: 1.320–7.958, p = 0.010) independently had a higher proportion of Gram-negative bacteria. The study reveals a high level of resistance to commonly used antimicrobials in our setting, necessitating strengthening of the antimicrobial stewardship program and routine surveillance of antimicrobial resistance patterns.

## Background

Sepsis is a global health problem and a leading cause of mortality and morbidity in patients admitted to the Intensive Care Unit. It has been declared by the World Health Organization (1). It has a significant contribution to an increase in mortality as well as an increase in health care costs, affecting around 48.9 million people each year (2). Resistance to commonly used antibiotics remains a major factor that complicates the management of sepsis worldwide (3).

Patients in the intensive care unit (ICU) are often at a high risk of sepsis. This is caused by their vulnerability, which is caused by a reduction in host defences and deregulation of immune responses. This is because of multiple procedures and the use of invasive devices, distorting the anatomical integrity and protective barriers, such as intubation, mechanical ventilation, central vascular access, and urinary catheters(4) (5).

Most of these patients have multiple comorbidities such as Diabetes Mellitus, cardiovascular diseases, hypertension, liver disease, renal disease, Human immunodeficiency virus/ acquired immunodeficiency syndrome [HIV/AIDS], which contribute to the risk of acquiring infection (6) (7).

In addition, several drugs routinely administered in the ICU are important risk factors for infection. For example, the use of sedatives and muscle relaxants reduces airway protective reflexes, leading to the accumulation of secretions. These secretions act as culture media for bacteria and cause respiratory tract infections(4) (8).

ICU infection leads to several adverse outcomes in patients, including increased morbidity and mortality, increased hospitalization costs, reduced health-related quality of life, and prolonged length of hospital stay (6).

Different research authors have demonstrated that the causes of sepsis in the ICU, in descending order, are lower respiratory tract infection, urinary tract infection, and catheter-related bloodstream infections (9)(10). The isolated bacteria were *Clostridium difficile, Klebsiella species, Acinetobacter baumanii, Enterococcus species, Pseudomonas aeruginosa, Citrobacter, Staphylococcus aureus, Staphylococcus pneumoniae, and Escherichia coli* (9) (11). Most Gram-positive isolates in LMICs were sensitive to vancomycin and linezolid, and gram-negative bacteria were sensitive to amikacin, imipenem, and tigecycline, with variable sensitivity to piperacillin-tazobactam, cefepime, and meropenem. There was also reported evidence of the presence of multidrug-resistant bacteria (MDR) (12)(13).

Patients’ age group was associated with the microbial profile in the ICU. Although gram-negative bacteria are the most common isolates in the ICU, the paediatric population demonstrates a predominance of gram-positive bacteria (14).

Currently, no study has identified the bacterial profiles and antibiotic sensitivity in the ICU, particularly in southern Tanzania. In addition, most studies lack a diverse population that includes a wide age range. This study will provide data covering a wide range of age groups. In addition, it will provide the bacterial profile and antibiotic sensitivity that fits our environment, which is important for managing sepsis, optimizing antimicrobial use, and improving patient outcomes. Understanding this profile positively influences treatment strategies, infection control measures, and overall patient safety.

## Methods

### Study design and setting

A descriptive and analytical prospective cross-sectional study was conducted at Mbeya Zonal Referral Hospital. The hospital is located in the southern highland zone of Tanzania. It operates as a southern zonal referral hospital serving five southern regions and as a teaching hospital under UDSM-MCHAS.

### Study duration

The study was conducted from April 2025 to March 2026

### Sample size and sampling technique

The sample size was obtained by using Kish and Lisle formula

The sample size for this study was 185.

A consecutive sampling technique was used, such that all patients who were eligible to participate during the study period were included in this study.

### Inclusion criteria

All sepsis-suspected patients in the ICU who were not on antibiotics or had been on antibiotics for less than 48 hours

### Data collection methods and procedures

Data were taken from Laboratory results of patients in the ICU, who were screened daily using the SIRS criteria to identify sepsis-suspected patients and whose specific samples were taken according to clinical judgement. All clinical specimens, including blood, urine, pus, CSF, sputum, and pleural fluid, were collected following standard sterile technique to prevent sample contamination (13). Blood samples were collected in two sterile culture bottles, one for aerobic bacteria and another for anaerobic bacteria in adults, and one bottle for paediatrics, which is aerobic. Other specimens were collected in sterile sample containers. Proper sample labelling and sample handling were done. Timely sample handover in the laboratory for analysis was done.

In the microbiology laboratory, collected samples were cultured under specific conditions. Starting with blood samples,

The culture bottles were kept in a BACTEC machine for five days maximum. The machine detects sample growth on any day, within five days. A preliminary Gram stain was conducted first, followed by inoculation of the sample into MacConkey agar, blood agar, and chocolate agar for 24 hours at 37 °C. Thereafter, the special biochemical tests were used to identify the isolates and confirm the specific species.

Urine, CSF, sputum, and pleural fluid samples were inoculated from sterile containers, pus samples were transported via Stuart transport media after being aseptically collected, and thereafter they were inoculated onto MacConkey agar and blood agar and cultured for 24 hours at 37 °C.

The isolated organisms from all samples were tested on their sensitivity patterns using the disc diffusion method, and the results were interpreted according to laboratory standards (16). Antibiotics tested in this study were amikacin, amoxiclav, ampicillin, azithromycin, ceftriaxone, cefepime, cefoxitin, ceftazidime, clindamycin, chloramphenicol, ciprofloxacin, gentamicin, imipenem, meropenem, piperacillin-tazobactam, trimethoprim-sulfamethoxazole, and tigecycline.

Data collection was done by the principal investigator, and two research assistants who were registrars in the department of anaesthesia and critical care at MZRH were trained on how to fill the checklist, as well as maintaining the confidentiality of the participants.

### Data analysis plan

Data collected in the study for bacterial profile and sensitivity pattern were analysed using SPSS version 27. Categorical variables were summarized using percentages and frequencies. These data include Age, sex, invasive devices, comorbidities, sample taken, Gram stain, isolated organism, and specific antibiotic sensitivity pattern.

Fisher’s exact test and chi-square test were used to show an association between patients’ demographics and microbial profile, as well as clinical characteristics and microbial profile.

Statistical significance was set at a p-value less than 0.05. Thereafter, modified Poisson regression was used in the univariate analysis to identify independent variables with a p-value of 0.25 as candidates for multivariate analysis to adjust for potential confounders. In the multivariate modified Poisson regression analysis, the p-value of 0.05 was considered statistically significant at 95% confidence interval.

### Ethical consideration

The ethical clearance was sought from the MUHAS Institutional Review Board (IRB) with authorization number MUHAS-REC-03-2025-2736, and authorization to conduct this research was obtained from the Executive Director at MZRH. Written informed consent was obtained from the study participants or their next of kin. For patients who cannot consent, a waiver was used to ask for consent on behalf of these patients. Confidentiality was ensured during the study. Also, participation in this study was fully voluntary, and participants or next of kin were allowed to withdraw from the study whenever they wanted

## RESULTS

### DEMOGRAPHIC AND CLINICAL CHARACTERISTICS

A total of 187 samples were included in this study, where the majority of samples were taken from patients in the age category of 15-35 years 63(33.7%). The study population was slightly male-predominant, with male patients accounting for 99(52.9%) of the patients. The most common primary admitting diagnosis was traumatic brain injury (TBI) (n = 48, 25.7%). Most patients had hypertension (n = 62, 43.7%). Most patients had a urinary catheter 176 (42.3%) (Table 1)

**Table 1:**
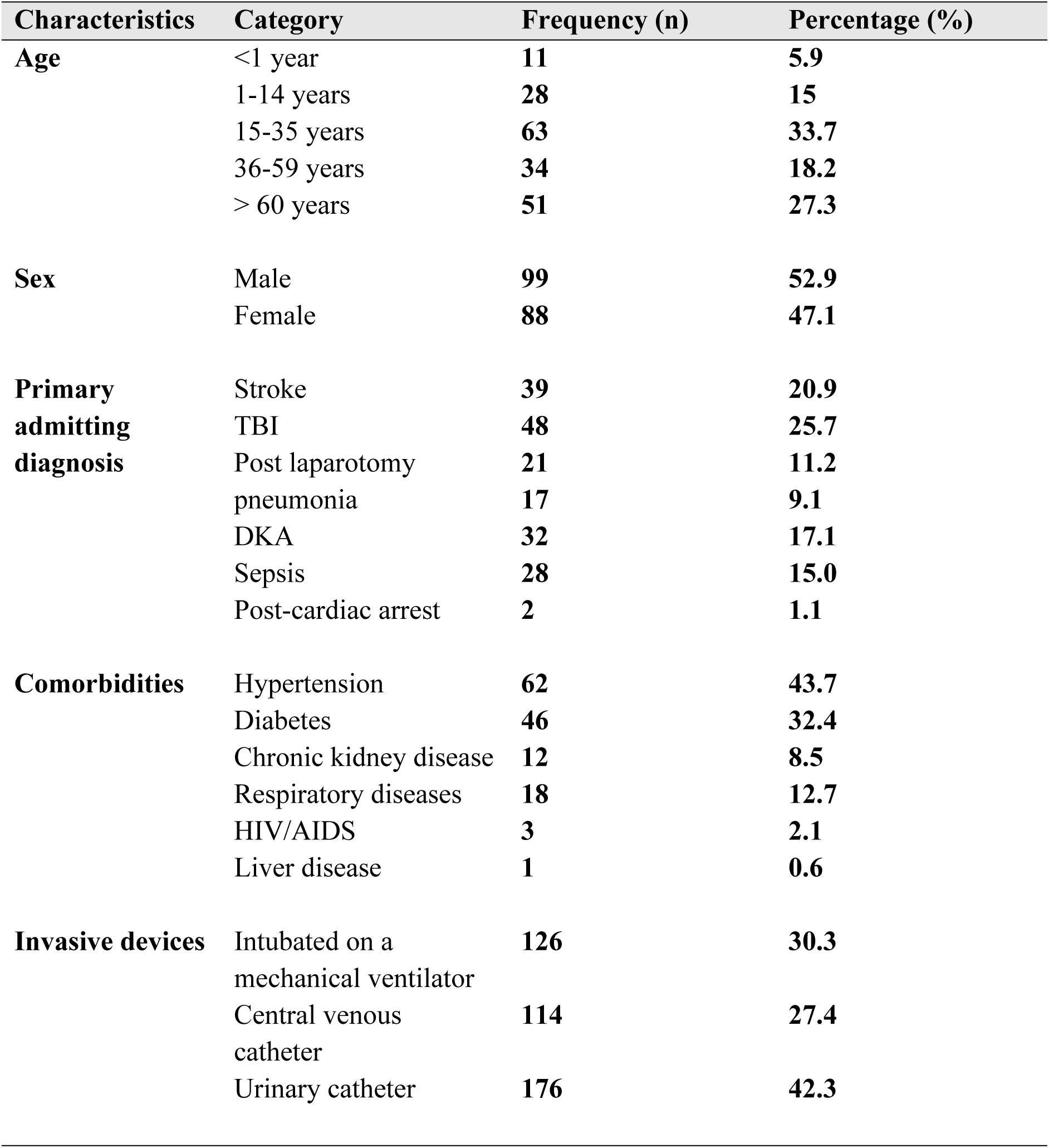
Distribution of Socio-Demographic and Clinical Characteristics of study participants (N=187)

### BACTERIAL PROFILE AMONG PATIENTS ADMITTED IN ICU

Of the 187 samples,78 (41.7%) were culture-positive. Among the culture-positive samples, gram-negative organisms were more common, accounting for 51(65.4%) of the samples. (Table 2).

**Table 2:**
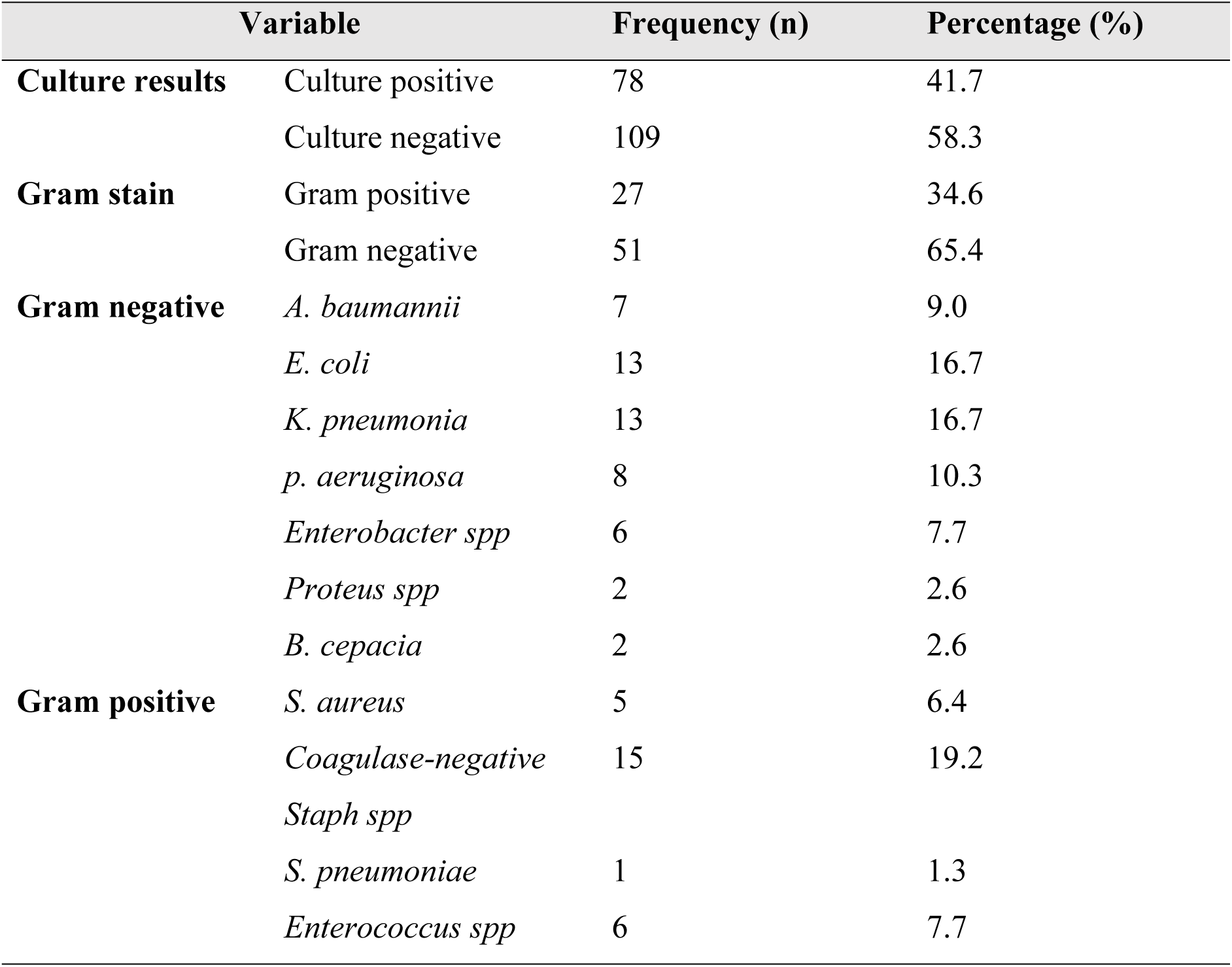
Bacterial profile isolated from ICU patients.

Among the Gram-negative isolates, *Escherichia coli* and *Klebsiella pneumoniae* were the most frequently identified, each contributing 13(16.7%) isolates. *Coagulase-negative Staphylococcus species* were the most common Gram-positive bacteria (n = 15, 19.2%). (Table 2).

In blood samples, *coagulase-negative Staphylococcus spp*. were commonly isolated (n = 15,34.1%). In pus samples, *E. coli* was frequently isolated 9 (47.4%). Urine samples had a predominance of *Enterococcus spp.* By 3(42.9%). Sputum and pleural fluid samples had a predominance of *K. pneumoniae*, with 6(100%) and 1(100%), respectively. The CSF sample contained only *P. aeruginosa* 1(100%). (Table 3)

**Table 3:** Distribution of isolated bacteria by specimen type (N =78)

| Bacterial isolate | Blood n<br>(%) | Urine n<br>(%) | Pus n<br>(%) | CSF n<br>(%) | Sputum<br>n (%) | Pleural<br>fluid n<br>(%) |
| --- | --- | --- | --- | --- | --- | --- |
| *A. baumannii* | 6 (13.6) | - | 1 (5.3) | - | - | - |
| *E. coli* | 2 (4.5) | 2<br>(28.6) | 9<br>(47.4) | - | - | - |
| *K. pneumonia* | 5 (11.4) | 1<br>(14.3) |  |  | 6(100) | 1 (100) |
| *S. aureus* | 5 (11.4) | - | - | - | - | - |
| *Coagulase-negative*<br>*Staph spp* | 15 (34.1) | - | - | - | - | - |
| *Strep. pneumonia* | 1 (2.3) | - | - | - | - | - |
| *p. aeruginosa* | 1 (2.3) | 1<br>(14.3) | 5<br>(26.3) | 1<br>(12.5) |  |  |
| *Enterobacter spp* | 5 (11.4) | - | 1 (5.3) | - | - | - |
| *Proteus spp* | - | - | 2<br>(10.5) | - | - | - |
| *enterococcus* | 2 (4.5) | 3<br>(42.9) | 1 (5.3) |  | - | - |
| *B. cepacia* | 2 (4.5) | - | - | - | - | - |

### ANTIBIOTICS SENSITIVITY PATTERN

#### ANTIBIOTICS SENSITIVITY PATTERN FOR GRAM-NEGATIVE BACTERIA

Among the tested bacteria, meropenem showed a relatively high sensitivity (49%), followed by imipenem (41.4%). Regarding the intermediate sensitivity pattern, imipenem had a high intermediate susceptibility rate (44.8%). Very high resistance rates were observed for ampicillin (100%). (Table 4).

**Table 4:**
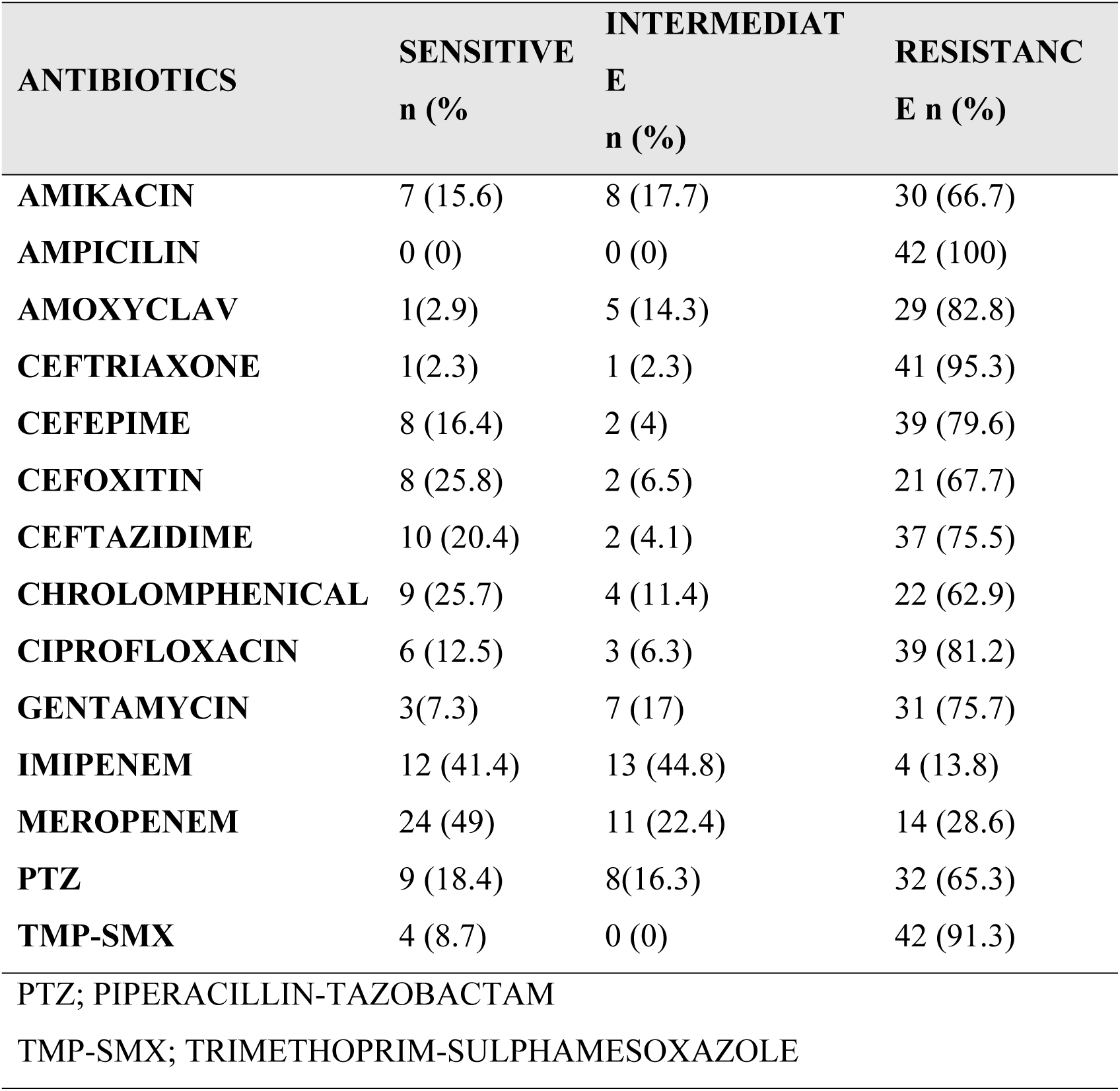
Description of antibiotic sensitivity patterns among ICU patients for Gram-negative bacteria.

#### ANTIBIOTIC SENSITIVITY PATTERN FOR GRAM-POSITIVE BACTERIA

Clindamycin exhibited the highest sensitivity (62.5%). A high intermediate pattern was observed with clindamycin (25% of isolates). High rates of azithromycin resistance were observed (94.1%). (Table 5).

**Table 5:** Description of antibiotic sensitivity pattern among ICU patients for Gram-positive bacteria.

| ANTIBIOTICS | SENSITIVE | INTERMEDIATE | RESISTANCE |
| --- | --- | --- | --- |
|  | n (%) | n (%) | n (%) |
| AMPICILIN | 1(11.1) | 1 (11.1) | 7(77.8) |
| AZITHROMYCIN | 1(5.9) | 0 (0) | 16 (94.1) |
| CEFOXITIN | 1 (5.9) | 1 (5.9) | 15 (88.2) |
| CHROLOMPHENICAL | 9 (45) | 4 (20) | 7 (35) |
| CIPROFLOXACIN | 5 (25) | 1 (5) | 14 (70) |
| CLINDAMYCIN | 5 (62.5) | 2 (25) | 1 (12.5) |
| GENTAMYCIN | 2 (9.5) | 0 (0) | 19 (90.5) |
| TRIMETHOPRIM-SULPHAMESOXAZOLE | 5 (25) | 0 (0) | 15 (75) |
| TIGECYCLINE | 3 (20) | 0 (0) | 12(80) |

**Table 6:**
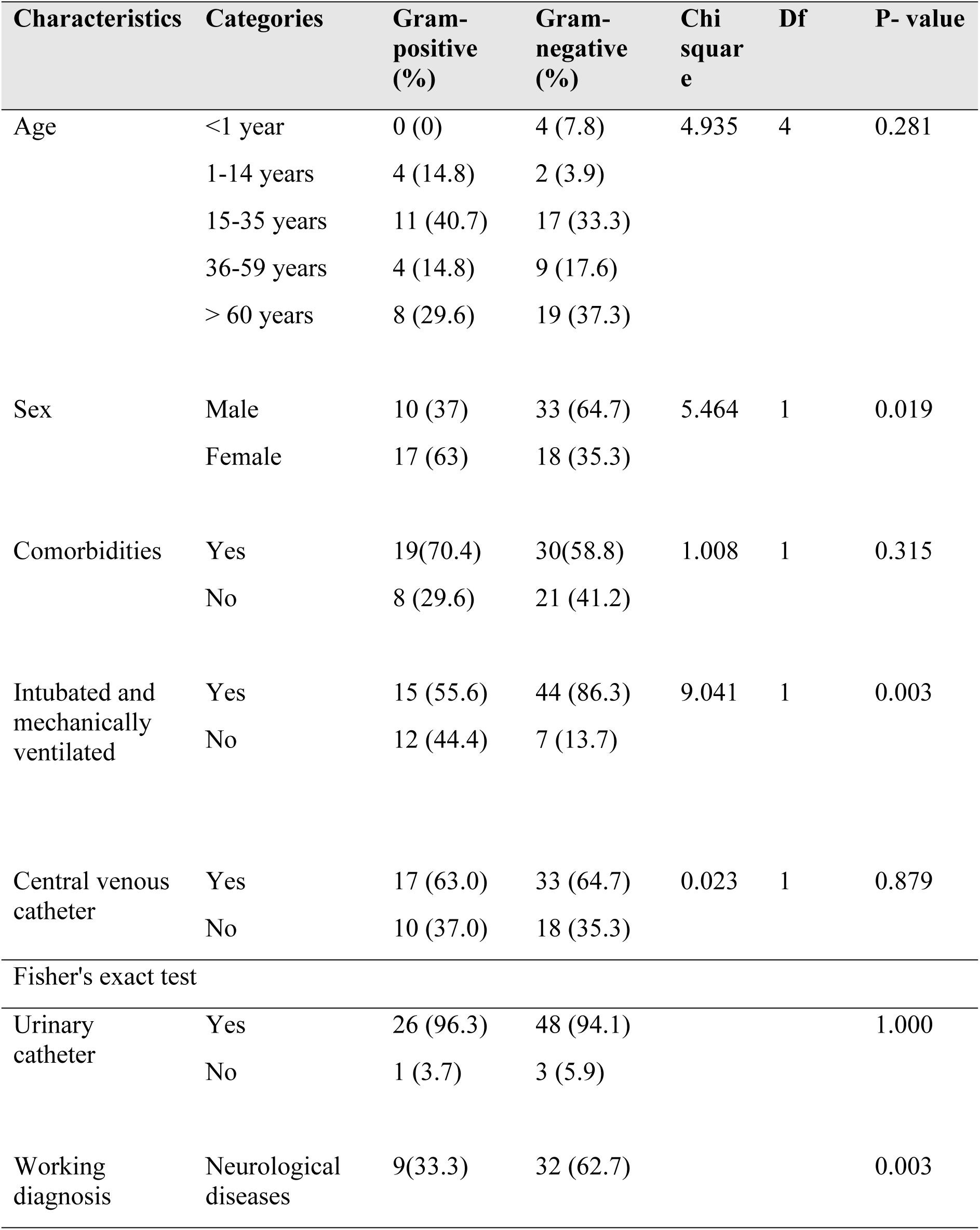
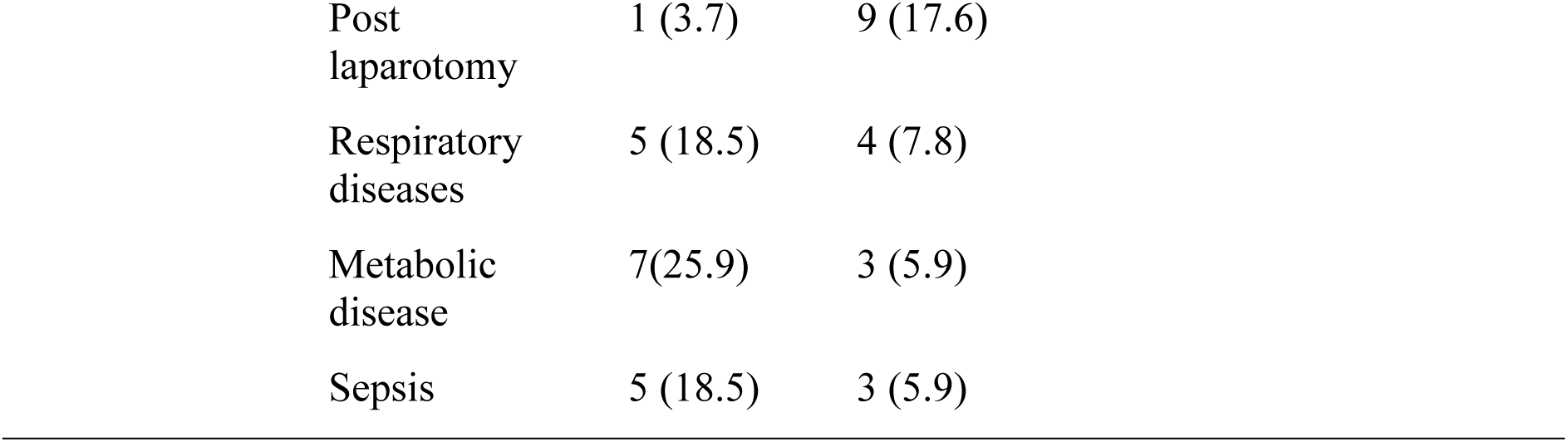
Association between patient characteristics and Gram stain of isolated bacteria (N=78)

**Table 7:** Univariate and Multivariate analysis (Modified Poisson Regression) of patient characteristics and Gram stain of isolated bacteria.

| Variable | Category | Crude<br>PR | 95%<br>CI | P<br>Value | APR | 95% CI | P-value |
| --- | --- | --- | --- | --- | --- | --- | --- |
| Sex | Male | 1.492 | 1.039-2.142 | 0.030 | 1.401 | 1.004-1.955 | 0.047 |
|  | Female | 1 |  |  | 1 |  |  |
| Working<br>diagnosis | Neurological<br>disease | 2.081 | 0.838-5.167 | 0.114 | 2.138 | 0.929-4.924 | 0.074 |
|  | Post<br>laparotomy | 2.400 | 0.958-6.011 | 0.062 | 3.532 | 1.516-8.228 | 0.003 |
|  | Respiratory<br>disease | 1.185 | 0.373-3.762 | 0.773 |  |  |  |
|  | Metabolic<br>disease | 0.800 | 0.217-2.943 | 0.737 |  |  |  |
|  | sepsis | 1 |  |  | 1 |  |  |
| ETT +<br>MV | Yes | 2.024 | 1.103-3.715 | 0.023 | 2.208 | 1.133-4.305 | 0.020 |
|  | No | 1 |  |  | 1 |  |  |

#### ASSOCIATION BETWEEN PATIENT DEMOGRAPHICS, CLINICAL CHARACTERISTICS, AND BACTERIAL PROFILE

The chi-square test showed that the sex of the patients (χ2 5.464, p = 0.019) and intubated and mechanically ventilated patients (χ2 9.041, p = 0.003) were statistically associated with the Gram stain of isolated bacteria. Working diagnosis (p = 0.003) showed a statistically significant association with Gram stain using Fisher’s exact test.

### UNIVARIATE AND MULTIVARIABLE MODIFIED POISSON REGRESSION ANALYSIS FOR ASSOCIATION BETWEEN PATIENT DEMOGRAPHICS, CLINICAL CHARACTERISTICS, AND BACTERIAL PROFILE

#### UNIVARIATE MODIFIED POISSON REGRESSION ANALYSIS

Univariate modified Poisson analysis showed that male patients (cPR 1.492, 95% CI 1.039-2.142, p 0.030), patients with neurologic diseases (cPR 2.081, 95% CI 0.838-5.167, p 0.114), patients who were post-laparotomy (cPR 2.400, 95% CI 0.958-6.011, p 0.062), and intubated and mechanically ventilated patients (cPR 2.024, 95% CI 1.103-3.715, p 0.023) had a high crude prevalence ratio of gram-negative bacteria.

### MULTIVARIATE MODIFIED POISSON REGRESSION ANALYSIS

In the multivariate modified Poisson analysis, after adjusting for possible confounders, male patients (aPR 1.401, 95% CI 1.004 - 1.955, p = 0.047), patients who were post-laparotomy patients (aPR 3.532, 95% CI 1.516 - 8.228, p = 0.003), and intubated and mechanically ventilated patients (aPR 2.208, 95% CI 1.113 - 4.305, p = 0.023) were independently associated with a higher adjusted prevalence ratio of having gram-negative bacteria.

## Discussion

### Bacterial profile among patients admitted to the ICU

More than half of the samples collected in this study were culture-negative. This finding could be due to the nature of patients in the ICU, where some present with sepsis-related symptoms while they are not septic. Some sepsis-causing organisms don’t grow in routine bacterial culture media. This finding mirrors the study conducted by Duan et al in China and Sharma et al in India (17).

Gram-negative bacteria predominated among those isolated in the current study. This finding is consistent with studies from both LMIC and developed countries, and it is due to comparable ICU-related risk factors such as the use of invasive medical devices and the ability of gram-negative bacteria to form biofilms on those devices, for example, ventilator tubing and catheters, thereby facilitating colonization, persistence, and transmission within the ICU environment.

### Antimicrobial sensitivity pattern among patients admitted to the ICU

#### Antimicrobial sensitivity for gram-negative bacteria

Gram-negative bacteria had considerable sensitivity towards carbapenems, with high resistance rates towards commonly used antibiotics, especially penicillin-based antibiotics and third- and fourth-generation cephalosporins. A Tigecycline evaluation surveillance trial also demonstrated similar findings among samples from the African Continent (18). However, the sensitivity of Gram-negative bacteria to amikacin and piperacillin-tazobactam, as reported in the TEST, was not reflected in our study. This difference may be due to differences in geographical locations and antimicrobial stewardship.

#### Antimicrobial sensitivity for gram-positive bacteria

Gram-positive isolates were relatively sensitive to clindamycin. This finding is similar to a study conducted in Canadian ICUs, which reported that some gram-positive bacteria, especially non-MRSA isolates, were sensitive to clindamycin (19). This similarity can be explained by the infrequent use of clindamycin in our study setting. However, these results differ from a study conducted in Iran, which portrayed a higher resistance rate of gram-positive bacteria to clindamycin(20). The author of the study in Iran highlighted the reasons for high clindamycin resistance, being its frequent usage in their ICU, as well as the presence of specific bacteria which had a high resistance rate towards clindamycin.

Furthermore, Gram-positive bacteria exhibited greater resistance to azithromycin, gentamicin, cefoxitin, and tigecycline. These results differ from studies done in China, where, in their study, gram-positive bacteria were sensitive to the mentioned antibiotics (17). The difference can be explained by differences in antimicrobial stewardship in the ICU setting and by geographical differences.

Another observation in this study is that the results highlighted the presence of multidrug-resistant bacteria in the ICU. This finding is similar to global data, where most ICUs have a higher proportion of MDR bacteria. The studies have shown the reasons for this finding to be inappropriate and excessive antimicrobial use, especially in the ICU setting; colonization of most of the ICU devices by multidrug-resistant bacteria; poor adherence to IPC protocols; and lack of local antimicrobial stewardship (4) (11) (21).

## ASSOCIATION BETWEEN PATIENT DEMOGRAPHICS, CLINICAL CHARACTERISTICS, AND BACTERIAL PROFILE

Male patients had a higher proportion of gram-negative bacteria than female patients. Our results were congruent with those of studies done in Ethiopia and Kenya (22) (23)which also demonstrated a predominance of gram-negative bacteria in male patients. This similarity may be due to the predominance of male patients in these studies. This finding differs from that reported in a study conducted in the UK, in which male patients demonstrated a predominance of gram-positive bacteria. The observed discrepancy may be attributed to differences in the study population, where the study in the UK involved patients with surgical site infection only (24). Nevertheless, male sex should not be interpreted as a risk factor for Gram-negative bacterial infection based on the present study, as the observed findings may have been influenced by the slight predominance of male participants in our study population.

In this study, patients who were intubated and mechanically ventilated had a higher proportion of Gram-negative bacteria. This finding aligns with a study done by Yusuph Savran in Turkey, a study done by EL-Tamimi in Jordan, and a study by Peter Agaba in Uganda (25) (26) (11). Intubated and mechanically ventilated patients are prone to prolonged ICU stay and immunosuppression secondary to critical illness, hence vulnerable to being colonized by ICU environment bacteria (4) (5). This serves as a reminder to all healthcare workers in the critical care unit to adhere to IPC protocols and regular ICU environment decontamination.

Patients who underwent post-laparotomy also had a higher proportion of gram-negative bacteria. This finding was similar to a study done by AL-Tamimi in Jordan Asia in the ICU, whereby they found that patients who were post-surgery had a higher chance of gram-negative bacterial infection in the ICU(26). These results are similar because patients in the perioperative period are vulnerable to infection secondary to the stress response to surgery. However, this finding differs from a study done by Pettini et al., where they found that patients from the medical ICU had a higher proportion of gram-negative bacteria(27). This difference could be attributed to the few patients enrolled in the surgical ICU in their study.

## STUDY LIMITATIONS

- Antibiotic testing was limited, especially for Gram-positive bacteria. This may lead to failure in generalizing the sensitivity patterns of isolated organisms. For example, sensitivity against vancomycin and linezolid was not tested in this study.
- Despite identifying the presence of multidrug-resistant bacteria, the inability to perform advanced diagnostic techniques, such as molecular testing, limited the ability to identify resistance mechanisms of isolated bacteria.
- This study was conducted at a single center, which may limit the ability to provide the epidemiology of different centers or different geographical locations.
- A cross-sectional study design doesn’t show a causal relationship.

## CONCLUSION AND RECOMMENDATION

### CONCLUSION

This study highlights the bacterial profile and antibiotic sensitivity patterns among patients admitted to the ICU at Mbeya Zonal Referral Hospital, with Gram-negative bacteria being commonly isolated. It demonstrates a high level of resistance to commonly used antibiotics, indicating that many first-line treatments are ineffective. Emerging carbapenem resistance is alarming, as carbapenems are the only antibiotics with a good sensitivity profile against Gram-negative bacteria in our setting. This poses a significant challenge to effective patient management. The findings of this study underscore the need for evidence-based antibiotic use and continuous microbiological surveillance.

### RECOMMENDATIONS

#### 1. CLINICAL PRACTICE RECOMMENDATIONS

- Revision of empirical antibiotic therapy: Given the high resistance rate to commonly used antibiotics, there is a need to revise local treatment guidelines in the ICU, whereby empirical antibiotic therapy should be guided by local antimicrobial susceptibility patterns.
- Rational use of antibiotics is important, and in this setting, carbapenems seem to be the last resort; however, there is an alarming emergence of resistance to them.

#### 2. HOSPITAL-LEVEL RECOMMENDATIONS

- Strengthening infection control and prevention measures, including regular environmental cleaning in the ICU and proper isolation protocols, is essential.
  - Including formation of an antimicrobial stewardship committee at the hospital level
- Adequate funding for the microbiology laboratory is required to ensure the availability of all required antibiotics for susceptibility testing, as well as molecular testing for resistance mechanisms of isolated bacteria.

#### 3. RESEARCH RECOMMENDATIONS

- Conducting further research on antibiotic stewardship in the ICU to identify factors associated with high resistance is recommended.

The implementation of the above recommendations will contribute to an understanding of the bacterial profile and antibiotic sensitivity pattern in the ICU setting, improvement of diagnostic approaches, and improvement of treatment to patients with sepsis in the ICU.

## Data Availability

The data underlying this study's findings come from laboratory results of samples collected from ICU patients who were suspected to have sepsis, who were not on antibiotics for more than 24 hours before sample collection, as part of routine care. Public deposition of the complete participant-level dataset is therefore restricted by the ethical approval and institutional data-governance requirements governing this study. The study was approved by the Muhimbili University of Health and Allied Sciences Research Ethics Committee (MUHAS-REC-03-2025-2736). De-identified data sufficient to reproduce the analyses reported in this article may be made available to qualified researchers following review and approval by the relevant institutional authorities and ethics committee, and subject to applicable data-sharing and data-transfer requirements. Access requests should be directed to the Muhimbili University of Health and Allied Sciences Research Ethics Committee or the appropriate institutional data-access authority at MUHAS. Data access is not controlled solely by the study authors.

## Acknowledgment

I extend my sincere appreciation to the Department of Anaesthesiology at Muhimbili University of Health and Allied Sciences and Mbeya Zonal Referral Hospital staff member

